# Men’s Willingness to Receive Text Messages and Talk with an HIV counselor from the National HIV Hotline in Tanzania for Support with Linkage to Care Following HIV self-testing

**DOI:** 10.1101/2024.06.01.24308312

**Authors:** Frank Mhando, Kelia Olughu, Marwa Nyankomo, James S. Ngocho, Ivan Teri, Gaspar Mbita, BRIDGE Africa Team, Donaldson F. Conserve

## Abstract

**Background:** Tanzania faces a significant burden of HIV, with particular challenges in reaching men and ensuring timely linkage to care. To address these issues, HIV self-testing (HIVST) has been implemented to increase HIV testing and the National HIV Hotlines are being considered as a strategy to facilitate linkage to care. This study aimed to assess the willingness of Tanzanian men to receive support from use the National HIV Hotline via mobile phones for HIVST and linkage to care.

**Methods:** Data from 505 men from the baseline survey of a cluster-randomized controlled trial conducted in June 2019 with 18 social networks or “camps” in Dar es Salaam, Tanzania. Participants were 18-year-old or older male camp members who were HIV-negative at the time of enrolment. Logistic regression models were used to assess factors associated with men’s comfort with talking with an HIV counselor over the phone.

**Results:** There were 505 heterosexual male participants enrolled in the study with an average age of 29 years. Logistic regression demonstrated that comfortability texting a friend about HIV self-testing (OR =3.37, 95% CI [1.97 – 5.76], being comfortable texting a friend about HIV (OR = 3.84, 95% CI [2.20 – 6.72], previous history of receiving HIV related text messages (aOR = 0.55, 95% CI [0.31 – 0.99] were significantly associated with men’s s comfortability talking to a HIV counselor on the National HIV Hotline following HIVST. The factors such as participants’ comfortability texting friend about HIVST (OR = 2.52, 95% CI [1.49 – 4.25]) and comfortability texting friend about HIV (OR = 2.96, 95% CI [1.83 – 4.80] were significantly associated with the probability of participant’s comfortability receiving text message from HIV counselor following HIVST.

**Conclusion:** These findings suggest an effort to develop and implement a user-friendly digital health intervention that promote comfortability, address private concerns, and deliver tailored support and information to individuals following HIV self-testing.

## Introduction

Tanzania has one of the highest burdens of HIV in the world, with an estimated 1.4 million people living with HIV in 2020 (1). The HIV prevalence among adults aged 15-64 is approximately 4.6%, and there were an estimated 36,000 new HIV infections in 2020 (2). HIV disproportionately affects certain populations in Tanzania, including women, young people, and key populations such as men who have sex with men, sex workers, and people who inject drugs (3). The government of Tanzania has made significant progress in its HIV response over the past decade, with increased access to HIV testing, treatment, and care services. As of 2020, approximately 82% of people living with HIV in Tanzania were aware of their status, and 82% of those who were aware of their status were receiving antiretroviral therapy (ART) (1,2). Additionally, the number of AIDS-related deaths has decreased by 43% since 2010 (2). Despite these gains, there are still significant challenges in the HIV response in Tanzania. Stigma and discrimination continue to be major barriers to testing, treatment, and care for people living with HIV, particularly for key populations. There are also challenges in reaching certain geographic areas and populations with HIV services, including rural areas and mobile populations.

In recent years there have been efforts to scale up innovative approaches to HIV testing, such as HIV self-testing, as a way to increase testing uptake and reach key populations (4–8). HIV self-testing (HIVST) has been shown to increase HIV testing uptake, particularly among populations that are hard to reach through traditional testing methods (8,9). HIVST has been shown to be acceptable and feasible in Tanzania, and is expected to increase HIV testing uptake (8,9).

HIVST was first introduced in Tanzania in 2017 and since then, the government has taken steps to promote its use as a means of increasing HIV testing uptake and improving access to testing among hard-to-reach populations (10). As of 2021, HIVST kits are available for purchase in pharmacies, and can also be distributed through community-based programs. The Ministry of Health, Community Development, Gender, Elderly, and Children (MOHCDGEC) has approved several HIVST kits for use in Tanzania, including OraQuick and Atomo, and has developed guidelines for their use. However, the use of HIVST is still relatively low in Tanzania compared to other testing methods. Also, challenges remain in ensuring that individuals who test positive through HIVST receive follow-up care and treatment, as well as addressing issues related to stigma and discrimination. Testing is only the first step in the HIV care continuum, and linkage to care following HIVST is crucial for achieving the UNAIDS 95-95-95 targets (3).

Studies have shown that digital health interventions, such as mobile phone text messaging or smartphone applications, have the potential to facilitate linkage to care by providing timely support, reminders, and information to individuals who have received positive HIVST results (11–15).

In Tanzania, the efforts to improve medication adherence among people living with HIV (PLHIV) have included the implementation of various digital health interventions. One notable approach involves the use of reminder short message services (SMS) texts, wherein PLHIV receive timely reminders to take their medication (16). Another intervention method explored is the real-time medication monitoring (RTMM) device (17), such as the Wisepill, which provides not only reminders but also allows the challenges of lifelong adherence to ART, which remain a significant issue for PLHIV in Tanzania and other resource limited settings. While SMS reminders and RTMM devices may offer valuable support, they do not necessarily lead to significant improvement in adherence rates among PLHIV in Kilimanjaro, Tanzania (18). Additionally, the National HIV Hotline has emerged as a vital resource in HIV prevention, providing people with access to information, counselling, and support services in Tanzania. Through the National HIV Hotline, individual can seek guidance on medication adherence, receive timely reminders, and access assistance in overcoming barriers to treatment adherence. However, it is not yet clear if HIVST users are willing to use the National HIV Hotline for support with linkage to care following HIV self-testing. This study aimed to investigate men’s willingness to use the National HIV Hotline for linkage to care services after HIVST.

## Material and Methods

### Data Source

We utilized data from the baseline survey of the STEP (**S**elf **T**esting **E**ducation **P**romotion) project, a five-year study funded by the National Institutes of Health (K99/R00MH110343) in Tanzania. The STEP project was conducted from 2018 in Dar es Salaam (the commercial capital of Tanzania and the city with the second-highest HIV prevalence) among heterosexual men aged 18 years and older (19). Participants of the STEP project were recruited from 18 camps (social networks comprising mainly young men) in the Manzese and Tandale wards of Dar es Salam, Tanzania (20). Participants were recruited in June 2019 with the following inclusion criteria: (1) male, (2) aged 18 years or older, (3) been a camp member for at least three months, and (4) self-report as HIV-negative at enrollment (21). Written informed consent was obtained from all participants prior to data collection. A total of 508 participants were screened; three participants did not meet eligibility criteria and 505 participants consented to participate into the study. The data used for analysis were accessed in April, 2023.

### Measures

The current study on men’s willingness to use the National HIV Hotline for support with linkage to care following HIVST has two outcomes’ variables; (1) comfortability talking with an HIV counselor on the National HIV hotline; and (2) comfortability receiving text messages from an HIV counselor. The outcome variables were measured with the questions *1) How comfortable would you feel talking with an HIV counselor on the National HIV hotline? And 2) How comfortable would you feel receiving text messages from an HIV counsel on your mobile phone?* Both questions were answered by 1) very comfortable 2) somewhat comfortable and 3) Uncomfortable. There were seven explanatory variables considered, including the participant’s role in the camp, history of receiving HIV-related text message, history of sending a text message to friend about HIV, comfortability texting close friend in camp about HIV, comfortability texting close friend about HIVST, willingness to receive information via mobile phone following HIVST, and comfortability sending self-test result via text.

### Statistical analysis

The main study outcomes focused on the comfortability of individuals in talking to a HIV Counselor on the National HIV Hotline following HIV self-testing (HIVST) and receiving text messages from a HIV Counselor after HIVST. Categorical variables were summarized using frequencies, percentages, and p-values to examine their correlation with the primary outcomes. Logistic regression analysis models were utilized to assess the associations between explanatory variables and the outcome of interest, presenting odds ratios along with confidence intervals. Variables with a significance level of p < 0.10 in bivariable analysis were included in the multivariable analysis. The multivariable logistic regression model was then applied to identify independent predictors of comfortability in talking to a HIV Counselor on the National HIV Hotline following HIVST and receiving text messages from a HIV Counselor after HIVST, with a significance level of p < 0.10. All statistical analyses were conducted using Stata statistical software, version 16.1. We described our categorical variables using the chi-square test, presenting frequencies, percentages, and p-values to assess the association between explanatory variables and our outcomes of interest. Logistic regression was then used for bivariate analysis, determining the likelihood of participant comfortability talking with an HIV counselor and comfortability receiving text messages from an HIV counselor. Significant variables (p < 0.10) were identified, and unadjusted odds ratios (ORs) were calculated. In the multivariate analysis, logistic regression explored factors affecting the comfortability of talking with an HIV counselor and the comfortability of receiving text messages from an HIV counselor. Variables with p < 0.10 from bivariate analysis were included. All selected variables were analyzed using multiple logistic regression, with significance set at p < 0.10. Adjusted odds ratios (aORs) were calculated to understand variable relationships. Stata statistical software, version 16.1, was utilized for all analyses.(22)

## Results

Our study had 505 participants, of which all were male, and most were single (63.8%), had secondary level education or greater (67.7%) and were in the age group 25 – 34 years (39.8%). Most of the participants had previously tested for HIV (90.3%), however, a majority had no history of using HIVST kit (91.5%). Regarding mobile phone access, the majority of the participants reported having a mobile phone (98.0%) and few participants reported sharing the mobile phone with another individual (19.4%). A few participants had a history of receiving HIV-related text message (28.5%) and sending a friend in camp text message about HIV (22.2%). The majority of participants indicated being comfortable texting friend about HIVST (82.8%) but a few indicated being comfortable texting friend about HIV (52.7%). Of the 505 participants, only 3.6% were aware of the National HIV Hotline. The majority of participants indicated being very comfortable talking to a HIV counselor on National HIV Hotline (84.2%) and receiving text from HIV Counselor (80.6%) following HIVST. Table 1 present the distribution of various characteristics and responses obtained from the study participants.

**Table 1:**
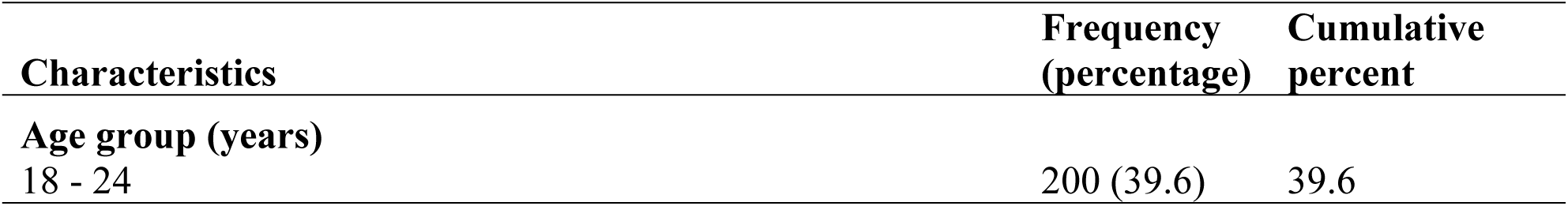
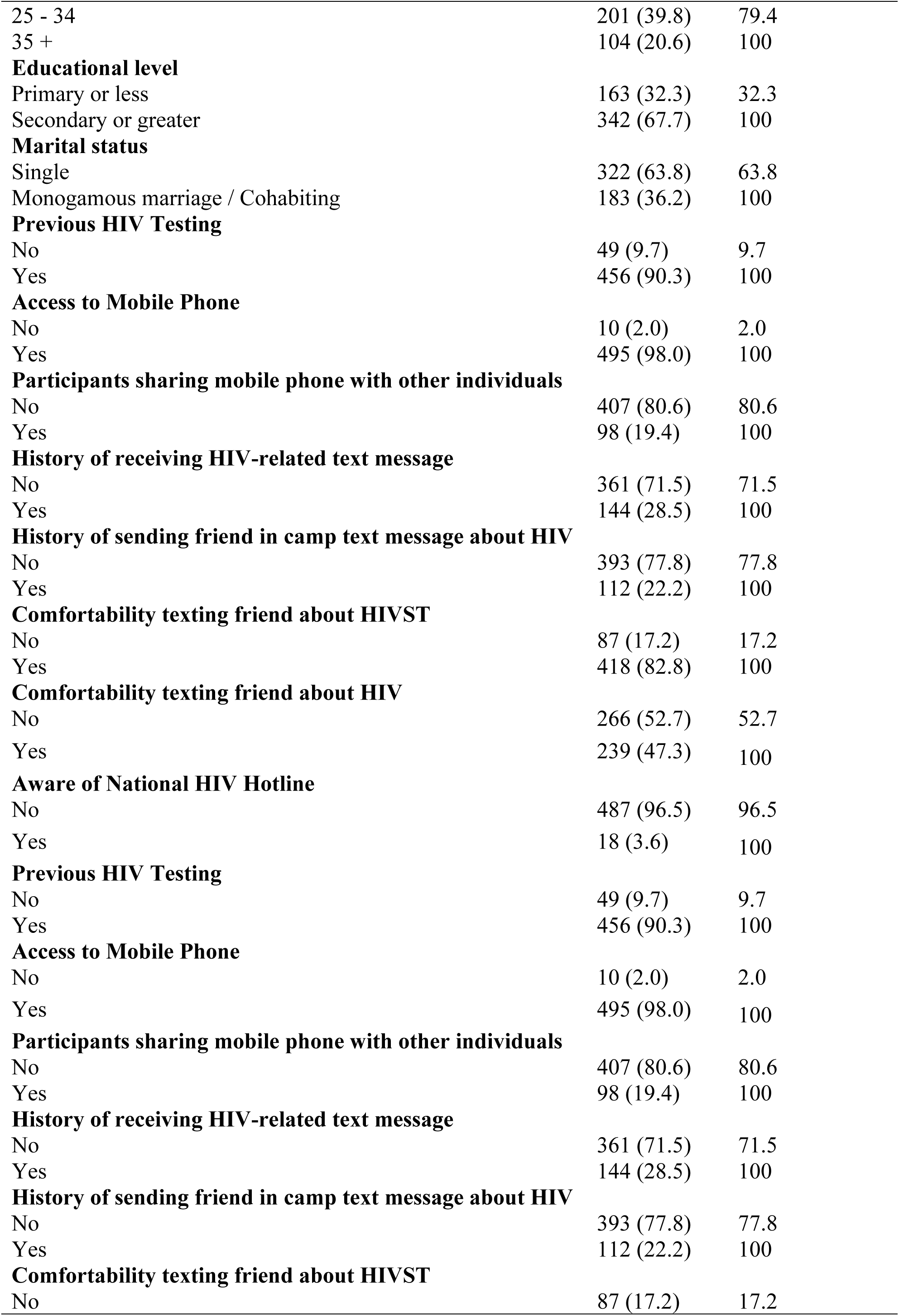
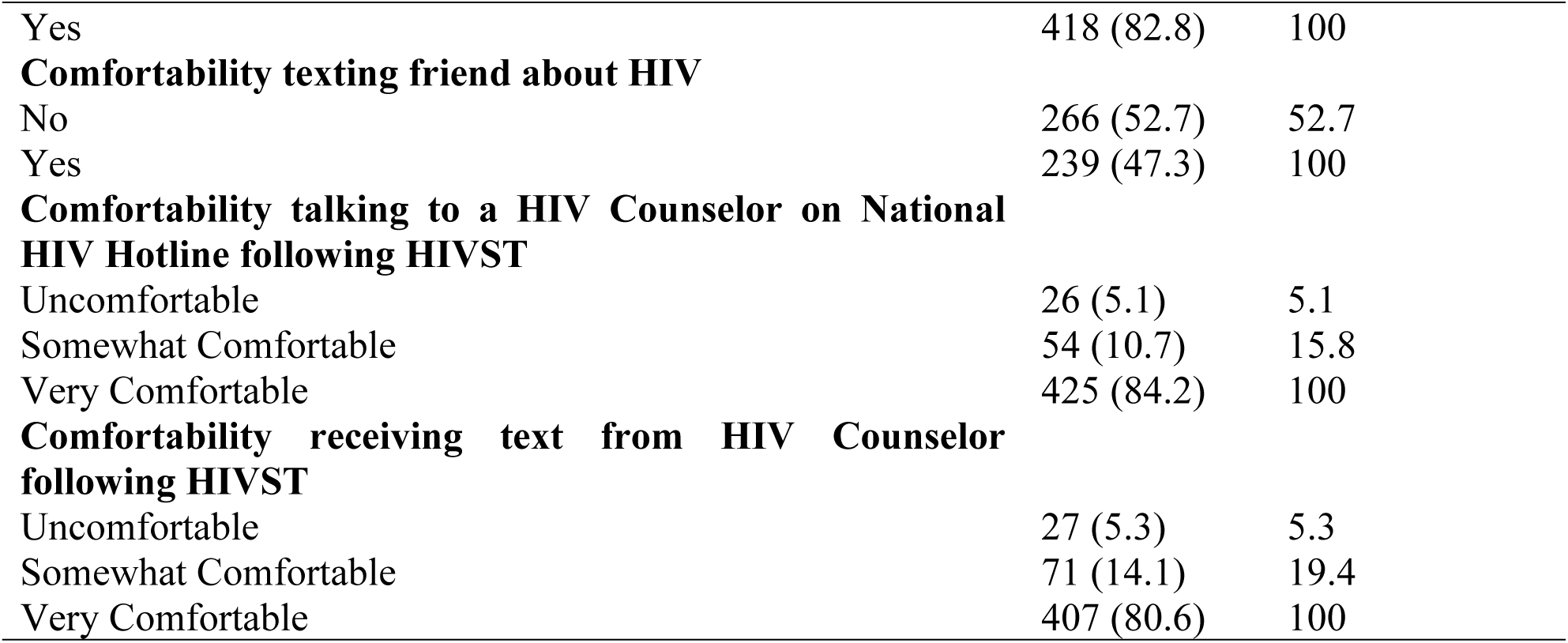
Participants’ characteristics, including demographic characteristics and HIV and HIVST knowledge and use. Dar es Salaam (n = 505)

| Characteristics | Frequency (percentage) | Cumulative percent |
| --- | --- | --- |
| Age group (years) |  |  |
| 18 - 24 | 200 (39.6) | 39.6 |
| 25 - 34 | 201 (39.8) | 79.4 |
| 35 + | 104 (20.6) | 100 |
| **Educational level** |  |  |
| Primary or less | 163 (32.3) | 32.3 |
| Secondary or greater | 342 (67.7) | 100 |
| **Marital status** |  |  |
| Single | 322 (63.8) | 63.8 |
| Monogamous marriage / Cohabiting | 183 (36.2) | 100 |
| **Previous HIV Testing** |  |  |
| No | 49 (9.7) | 9.7 |
| Yes | 456 (90.3) | 100 |
| **Access to Mobile Phone** |  |  |
| No | 10 (2.0) | 2.0 |
| Yes | 495 (98.0) | 100 |
| **Participants sharing mobile phone with other individuals** |  |  |
| No | 407 (80.6) | 80.6 |
| Yes | 98 (19.4) | 100 |
| **History of receiving HIV-related text message** |  |  |
| No | 361 (71.5) | 71.5 |
| Yes | 144 (28.5) | 100 |
| **History of sending friend in camp text message about HIV** |  |  |
| No | 393 (77.8) | 77.8 |
| Yes | 112 (22.2) | 100 |
| **Comfortability texting friend about HIVST** |  |  |
| No | 87 (17.2) | 17.2 |
| Yes | 418 (82.8) | 100 |
| **Comfortability texting friend about HIV** |  |  |
| No | 266 (52.7) | 52.7 |
| Yes | 239 (47.3) | 100 |
| **Aware of National HIV Hotline** |  |  |
| No | 487 (96.5) | 96.5 |
| Yes | 18 (3.6) | 100 |
| **Previous HIV Testing** |  |  |
| No | 49 (9.7) | 9.7 |
| Yes | 456 (90.3) | 100 |
| **Access to Mobile Phone** |  |  |
| No | 10 (2.0) | 2.0 |
| Yes | 495 (98.0) | 100 |
| **Participants sharing mobile phone with other individuals** |  |  |
| No | 407 (80.6) | 80.6 |
| Yes | 98 (19.4) | 100 |
| **History of receiving HIV-related text message** |  |  |
| No | 361 (71.5) | 71.5 |
| Yes | 144 (28.5) | 100 |
| **History of sending friend in camp text message about HIV** |  |  |
| No | 393 (77.8) | 77.8 |
| Yes | 112 (22.2) | 100 |
| **Comfortability texting friend about HIVST** |  |  |
| No | 87 (17.2) | 17.2 |
| Yes | 418 (82.8) | 100 |
| **Comfortability texting friend about HIV** |  |  |
| No | 266 (52.7) | 52.7 |
| Yes | 239 (47.3) | 100 |
| **Comfortability talking to a HIV Counselor on National HIV Hotline following HIVST** |  |  |
| Uncomfortable | 26 (5.1) | 5.1 |
| Somewhat Comfortable | 54 (10.7) | 15.8 |
| Very Comfortable | 425 (84.2) | 100 |
| **Comfortability receiving text from HIV Counselor following HIVST** |  |  |
| Uncomfortable | 27 (5.3) | 5.3 |
| Somewhat Comfortable | 71 (14.1) | 19.4 |
| Very Comfortable | 407 (80.6) | 100 |

### Comfortability talking to a HIV Counselor on National HIV Hotline following HIVST

In the bivariate analysis, we found that comfortability texting friend about HIVST (OR = 3.37, 95% CI [1.97 – 5.76]) and comfortability texting friend about HIV (OR = 3.84, 95% CI [2.20 – 6.72]) were significantly associated with the probability of participant’s comfortability talking to a HIV counselor on National HIV Hotline following HIVST (Table 2).

**Table 2:**
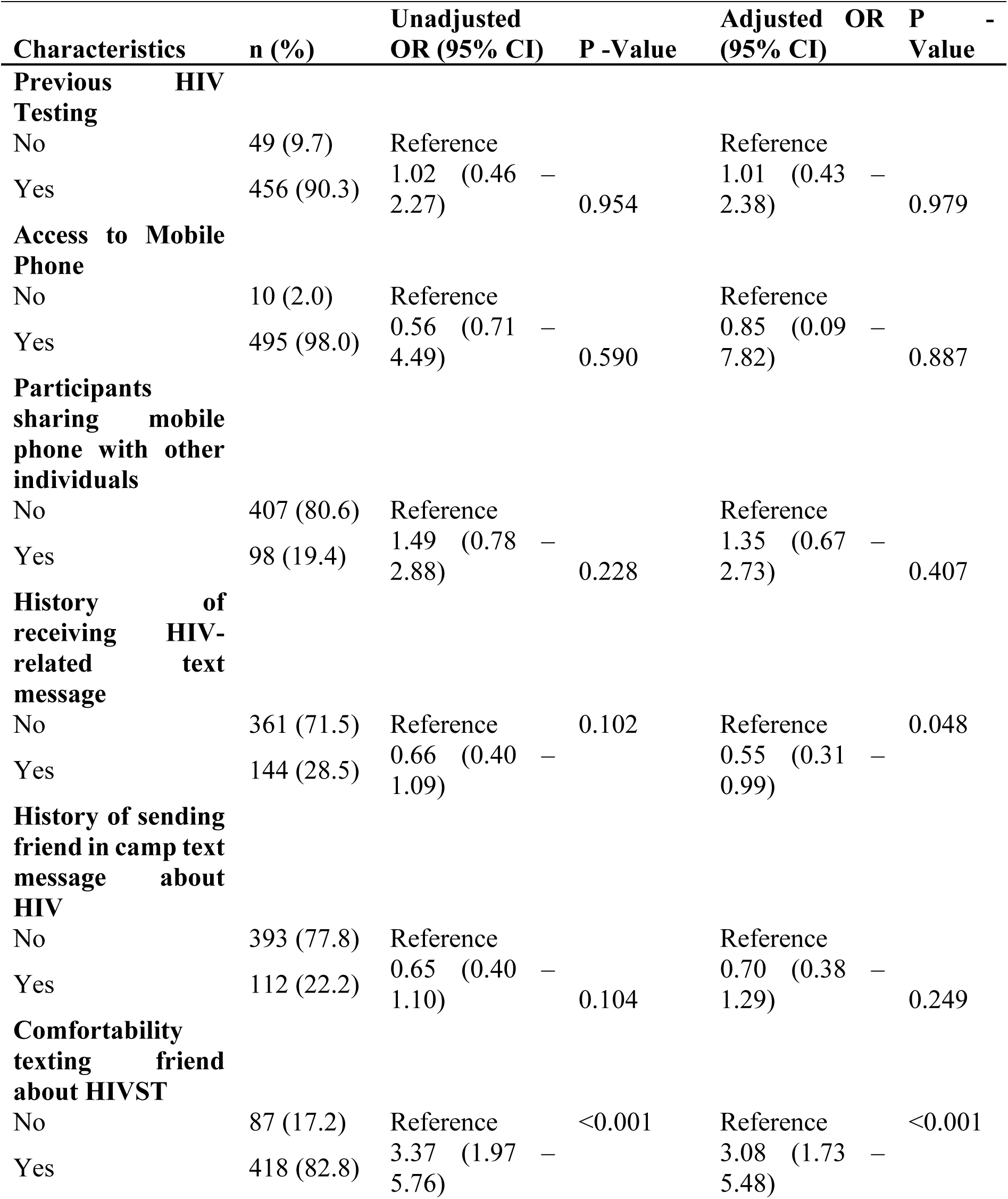
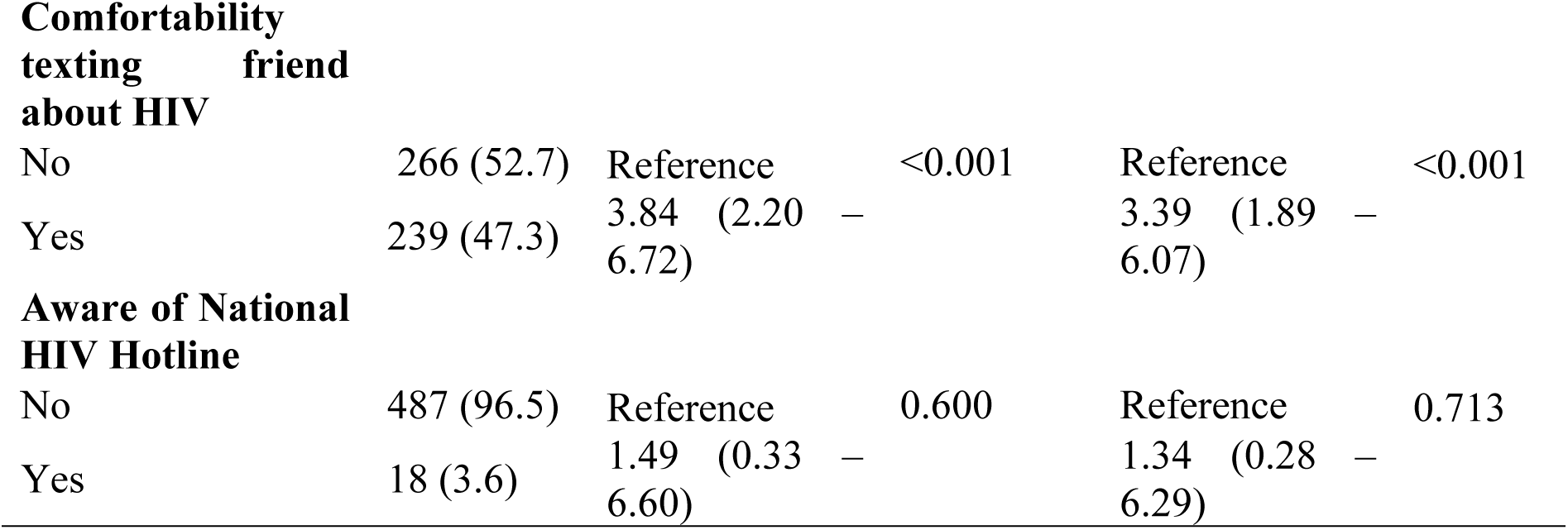
Logistic regression of the association between participants’ comfortability talking to a HIV Counselor on National HIV Hotline following HIVST and predictor variables showing unadjusted and adjusted odds ratio.

| Characteristics | n (%) | Unadjusted OR (95% CI) | P -Value | Adjusted OR (95% CI) | P -Value |
| --- | --- | --- | --- | --- | --- |
| **Previous HIV Testing** |  |  |  |  |  |
| No | 49 (9.7) | Reference |  | Reference |  |
| Yes | 456 (90.3) | 1.02 (0.46 – 2.27) | 0.954 | 1.01 (0.43 – 2.38) | 0.979 |
| **Access to Mobile Phone** |  |  |  |  |  |
| No | 10 (2.0) | Reference |  | Reference |  |
| Yes | 495 (98.0) | 0.56 (0.71 – 4.49) | 0.590 | 0.85 (0.09 – 7.82) | 0.887 |
| **Participants sharing mobile phone with other individuals** |  |  |  |  |  |
| No | 407 (80.6) | Reference |  | Reference |  |
| Yes | 98 (19.4) | 1.49 (0.78 – 2.88) | 0.228 | 1.35 (0.67 – 2.73) | 0.407 |
| **History of receiving HIV-related text message** |  |  |  |  |  |
| No | 361 (71.5) | Reference | 0.102 | Reference | 0.048 |
| Yes | 144 (28.5) | 0.66 (0.40 – 1.09) |  | 0.55 (0.31 – 0.99) |  |
| **History of sending friend in camp text message about HIV** |  |  |  |  |  |
| No | 393 (77.8) | Reference |  | Reference |  |
| Yes | 112 (22.2) | 0.65 (0.40 – 1.10) | 0.104 | 0.70 (0.38 – 1.29) | 0.249 |
| **Comfortability texting friend about HIVST** |  |  |  |  |  |
| No | 87 (17.2) | Reference | <0.001 | Reference | <0.001 |
| Yes | 418 (82.8) | 3.37 (1.97 – 5.76) |  | 3.08 (1.73 – 5.48) |  |
| **Comfortability texting friend about HIV** |  |  |  |  |  |
| No | 266 (52.7) | Reference | <0.001 | Reference | <0.001 |
| Yes | 239 (47.3) | 3.84 (2.20 – 6.72) |  | 3.39 (1.89 – 6.07) |  |
| **Aware of National HIV Hotline** |  |  |  |  |  |
| No | 487 (96.5) | Reference | 0.600 | Reference | 0.713 |
| Yes | 18 (3.6) | 1.49 (0.33 – 6.60) |  | 1.34 (0.28 – 6.29) |  |

In the multivariate analysis, after for the other predictor variables, we found that history of receiving HIV-related text message, comfortability texting friend about HIVST, and comfortability texting friend about HIV remained significantly associated with the probability of participant’s comfortability talking to a HIV counselor on National HIV Hotline following HIVST (Table 2). Participants with a previous history of receiving HIV related text message had 0.55 higher odds of being very comfortable talking to a HIV counselor on National HIV Hotline following HIVST (aOR = 0.55, 95% CI [0.31 – 0.99]). Participants who reported being comfortable texting a friend about HIVST had 3.08 higher odds of being very comfortable talking to a HIV counselor on National HIV Hotline following HIVST (aOR = 3.08, 95% CI [1.73 – 5.48]). Finally, participants who reported being comfortable texting friend about HIV had 3.39 higher odds of being very comfortable talking to a HIV counselor on National HIV Hotline following HIVST (aOR = 3.39, 95% CI [1.89 – 6.07]).

### Comfortability receiving text message from HIV Counselor following HIVST

In the bivariate analysis, we found that comfortability texting friend about HIVST (OR = 2.52, 95% CI [1.49 – 4.25]) and comfortability texting friend about HIV (OR = 2.96, 95% CI [1.83 – 4.80]) were significantly associated with the probability of participant’s comfortability receiving text message from HIV counselor following HIVST (Table 3).

**Table 3:**
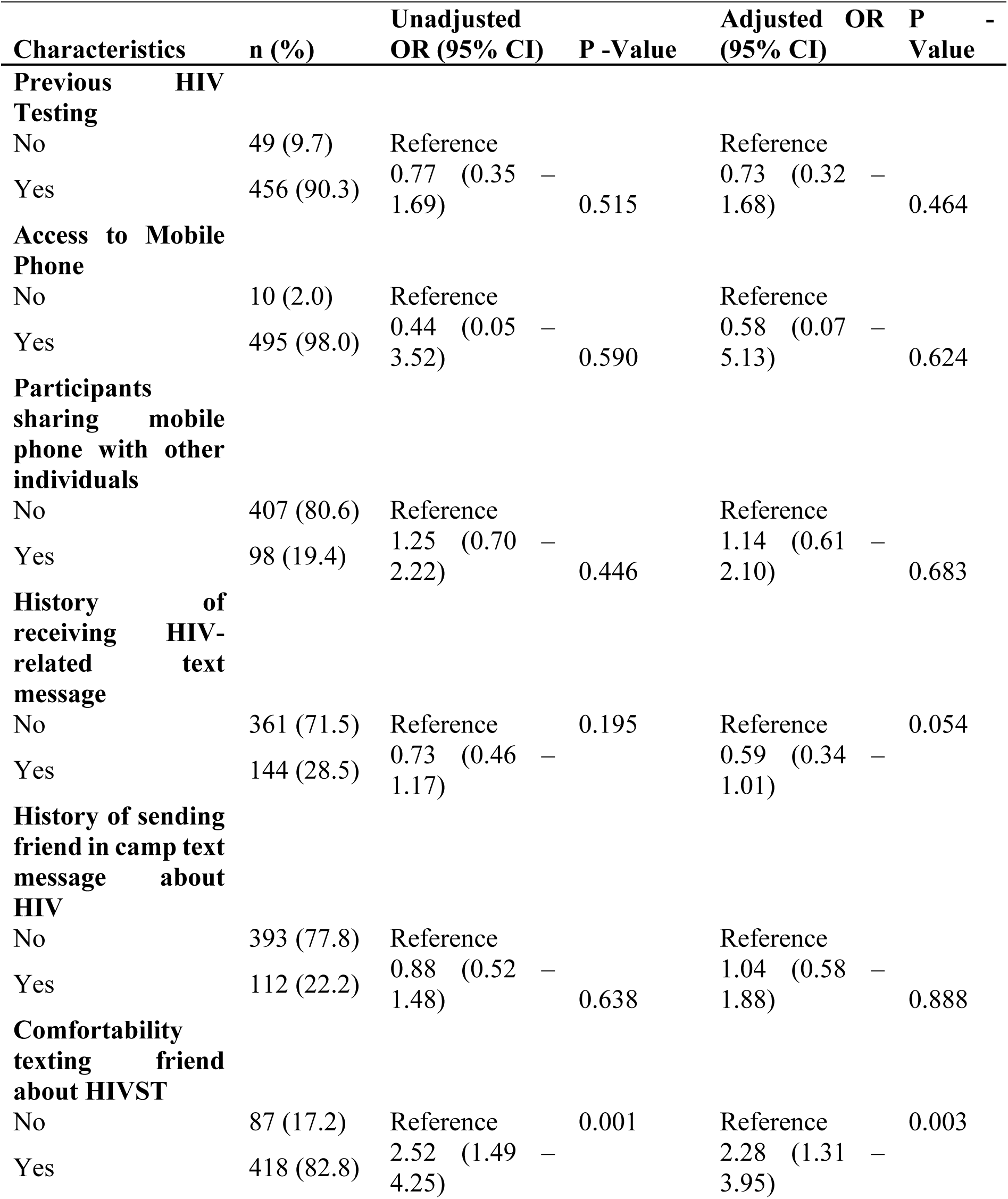
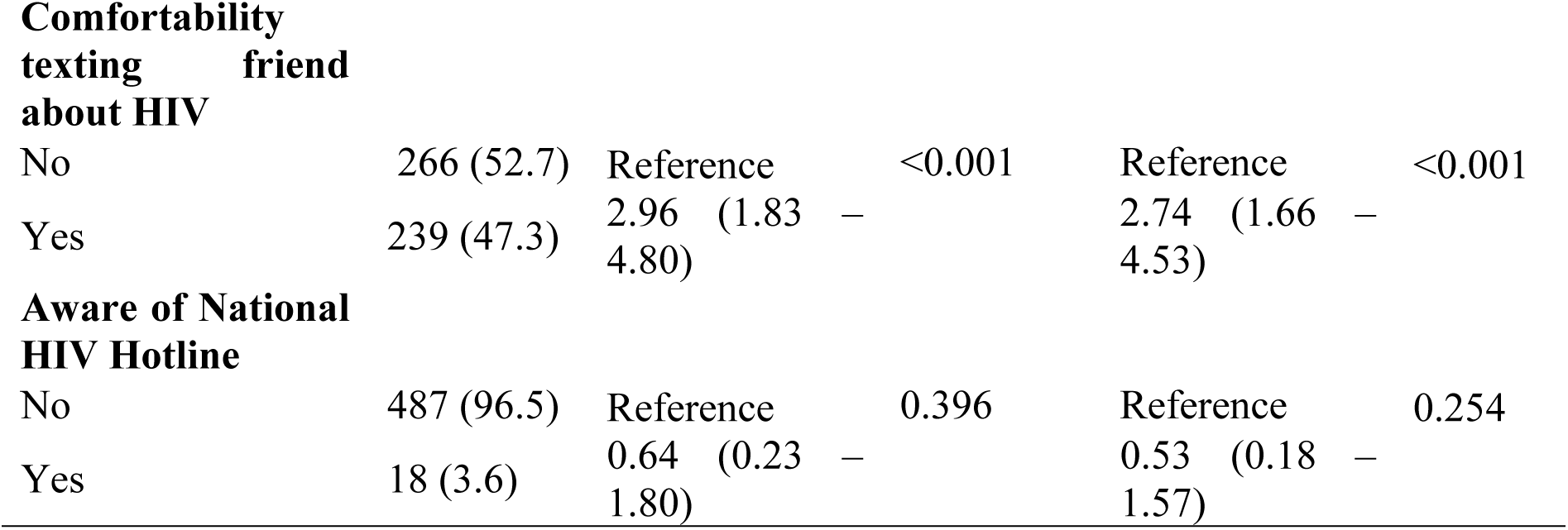
Logistic regression of the association between participants’ comfortability receiving text from HIV Counselor following HIVST and predictor variables showing unadjusted and adjusted odds ratio.

| Characteristics | n (%) | Unadjusted OR (95% CI) | P -Value | Adjusted OR (95% CI) | P -Value |
| --- | --- | --- | --- | --- | --- |
| **Previous HIV Testing** |  |  |  |  |  |
| No | 49 (9.7) | Reference |  | Reference |  |
| Yes | 456 (90.3) | 0.77 (0.35 – 1.69) | 0.515 | 0.73 (0.32 – 1.68) | 0.464 |
| **Access to Mobile Phone** |  |  |  |  |  |
| No | 10 (2.0) | Reference |  | Reference |  |
| Yes | 495 (98.0) | 0.44 (0.05 – 3.52) | 0.590 | 0.58 (0.07 – 5.13) | 0.624 |
| **Participants sharing mobile phone with other individuals** |  |  |  |  |  |
| No | 407 (80.6) | Reference |  | Reference |  |
| Yes | 98 (19.4) | 1.25 (0.70 – 2.22) | 0.446 | 1.14 (0.61 – 2.10) | 0.683 |
| **History of receiving HIV-related text message** |  |  |  |  |  |
| No | 361 (71.5) | Reference | 0.195 | Reference | 0.054 |
| Yes | 144 (28.5) | 0.73 (0.46 – 1.17) |  | 0.59 (0.34 – 1.01) |  |
| **History of sending friend in camp text message about HIV** |  |  |  |  |  |
| No | 393 (77.8) | Reference |  | Reference |  |
| Yes | 112 (22.2) | 0.88 (0.52 – 1.48) | 0.638 | 1.04 (0.58 – 1.88) | 0.888 |
| **Comfortability texting friend about HIVST** |  |  |  |  |  |
| No | 87 (17.2) | Reference | 0.001 | Reference | 0.003 |
| Yes | 418 (82.8) | 2.52 (1.49 – 4.25) |  | 2.28 (1.31 – 3.95) |  |
| **Comfortability texting friend about HIV** |  |  |  |  |  |
| No | 266 (52.7) | Reference | <0.001 | Reference | <0.001 |
| Yes | 239 (47.3) | 2.96 (1.83 – 4.80) |  | 2.74 (1.66 – 4.53) |  |
| **Aware of National HIV Hotline** |  |  |  |  |  |
| No | 487 (96.5) | Reference | 0.396 | Reference | 0.254 |
| Yes | 18 (3.6) | 0.64 (0.23 – 1.80) |  | 0.53 (0.18 – 1.57) |  |

In the multivariate analysis, after for the other predictor variables, we found that history of receiving HIV-related text message, comfortability texting friend about HIVST, and comfortability texting friend about HIV remained significantly associated with the probability of participant’s comfortability talking to a HIV counselor on National HIV Hotline following HIVST (Table 2). Participants with a previous history of receiving HIV related text message had 0.59 higher odds of being very comfortable talking to a HIV counselor on National HIV Hotline following HIVST (aOR = 0.59, 95% CI [0.34 – 1.01]). Participants who reported being comfortable texting a friend about HIVST had 2.28 higher odds of being very comfortable talking to a HIV counselor on National HIV Hotline following HIVST (aOR = 2.28, 95% CI [1.31 – 3.95]). Finally, participants who reported being comfortable texting friend about HIV had 2.74 higher odds of being very comfortable talking to a HIV counselor on National HIV Hotline following HIVST (aOR = 2.74, 95% CI [1.66 – 4.53]).

## Discussion

The overall goal of this study was to assess men’s willingness to use the National HIV Hotlines for support with linkage to care following HIV self-testing results in Tanzania. The results revealed several significant findings. A vast majority of participants reported having access to mobile phones (98.0%), indicating potential feasibility for utilizing mobile health (mHealth) interventions such as text messaging for HIV-related communication. These findings align with and reinforce the insights gleaned from previous research conducted in resource-limited setting (23–27). The studies have consistently demonstrated the extensive use of mobile phones even in regions with limited access to traditional healthcare infrastructure. The ubiquity of mobile phones in such setting has been pivotal in reaching remote and underserved populations with healthcare information and services.

Regarding the comfortability in engaging with HIV counseling services, participants who reported receiving HIV-related text messages in the past were more likely to feel comfortable talking to a HIV counselor on the National HIV Hotline following HIVST. This underscores the potential influence of previous exposure to HIV-related information and communication on individuals’ comfort levels in seeking HIV-related services. It also highlights the importance of targeted messaging and communication strategies in promoting awareness and comfortability with HIV testing and counseling services.

Among our study population, the majority of participants (82.77%) expressed comfort in texting friends about HIVST, which suggests a high level of acceptance and openness towards discussing HIV-related topics among peers. This finding aligns with previous research highlighting the importance of social support networks in HIV prevention and care. Peer influence has been recognized as a cornerstone of effective HIV interventions, Peers often individuals who share similar life experiences and challenges, can offer unique insights, empathy, and encouragement. They can play a pivotal role in motivating individuals to take control of their health and seek essential services, including HIV testing and care (28). Also, willingness to discuss HIV self-testing with close friends can contribute to reducing stigma associated with HIV. Peer support can create a sense of belonging and reduce isolation that some individuals living with HIV may experience (29,30)

The study’s findings also emphasize the importance of tailored communication and messaging strategies in promoting comfortability and uptake of HIV-related services, particularly among young, male populations in Tanzania. By leveraging mobile phone technology and peer networks, mHealth interventions could play a crucial role in enhancing access to HIV testing, counseling, and support services, thereby contributing to HIV prevention and care efforts (31). Thus, although this study focused on Men’s willingness to use the National HIV Hotlines for support with linkage to care following HIV self-testing results in Tanzania, there is benefit in future study investigating both sexual partner’s willingness to use the digital health intervention for HIVST and linkage to care services. As future study could offer additional insight into whether prior discussion of HIVST with sexual partner could be a potential explanation for the proportion of willingness to use digital health intervention among heterosexual men and their sexual partners.

Although our study had numerous strengths, there were a few limitations. First, the data originated from a cross-sectional survey and cannot be utilized in assessing the causal relationship between the variables of interest. Second, the study’s overall objective is to inform the potential for digital health intervention to support HIVST and linkage to care services however, this study collected data solely from male participants and lacked information on respondents’ sexual partners and their willingness to use digital health intervention for HIVST. Another limitation was the lack of sexual behavior data, as although the study gathered information on number of sexual partners within 12 months for each respondent, it did not include data on sexual behavior for analysis, such as condom use or PrEP use. Finally, the study findings are not generalizable to other men in Tanzania as participants were recruited among camps (social networks). Although not generalizable, this is one of few studies to assess partner-delivered HIVST kit distribution potential among men exposed to peer-led HIV self-testing education and promotion.

## Conclusion

Our study sheds light on men’s willingness to utilize the National HIV Hotlines for support with linkage to care following HIV self-testing results in Tanzania. The findings underscore the potential feasibility and acceptance of mobile health (mHealth) interventions, particularly text messaging, in facilitating HIV-related communication and service uptake among men in resource-limited settings. The ubiquity of mobile phones among participants highlights the opportunity to leverage digital health technologies to reach remote and underserved populations with HIV testing, counseling, and support services. Moreover, our findings emphasize the influence of previous exposure to HIV-related information and communication on individuals’ comfort levels in seeking HIV counseling services, indicating the importance of targeted messaging strategies. Furthermore, the high level of acceptance and openness towards discussing HIV-related topics among peers underscores the significance of peer support networks in HIV prevention and care. Leveraging peer influence and social networks can contribute to reducing stigma associated with HIV, promoting HIV testing uptake, and facilitating linkage to care.

However, the study has limitations, the focus solely on male participants limits the generalizability of the findings to other populations, particularly sexual partners. Future research should explore both partners’ willingness to use digital health interventions for HIV self-testing and linkage to care services to provide a more comprehensive understanding of the potential impact of such interventions.

Despite these limitations, our study contributes to the growing body of evidence on the feasibility and acceptability of the National HIV Hotlines for HIV testing and linkage to care in resource-limited settings. By addressing barriers to HIV service uptake and leveraging digital health technologies, we can advance HIV prevention and care efforts and work towards achieving the goals of HIV epidemic control.

## Data Availability

Please note that the data underlying this study will be made available upon special request

## Abbreviations

AIDS: Acquired Immune Deficiency Syndrome
HIV: Human Immunodeficiency Virus
HIVST: HIV self-testing
STEP: Self-Testing Education and Promotion
mHealth: Mobile Health

## Acknowledgement and Declarations

Funding: This study was supported by grants from the National Institute of Mental Health (Grant #R00MH110343: PI: DFC) and Minority Health International Research Training (MHIRT) (Grant T37-MD001448) from the National Institute on Minority Health and Health Disparities, National Institutes of Health (NIH), Bethesda, MD, USA.

## Competing Interests

No competing of interests

## Ethics approval and consent to participate

This study utilized secondary data collected from the previous STEP project, which was implemented starting in 2018. The initial ethics approval for that project was obtained from the University of South Carolina, covering the period from May 2017 to May 2018, during which data collection took place. The project received approval from different institutions due to the Principal Investigator (PI) shifting institutions during the project. Specifically, the project was reviewed and approved by the institutional review boards of the University of South Carolina approval period: 5/16/2017 to 5/15/2018), the National Institute of Medical Research of Tanzania (NIMR/HQ/R.8c/Vol.I/1170, approval period: 29th June 2017 to 28th June 2018), and George Washington University in 2021. All respondents provided written informed consent prior to participation in the original study. All study procedures were performed in accordance with the ethical standards of the institutional and/or national research committees and the 1964 Helsinki Declaration and its later amendments or comparable ethical standards.

For our current study, which involves secondary analysis of this already collected data, we did not seek an extension of the original ethics approval. According to the guidelines of our institutional review boards and standard ethical practices, secondary data analysis does not typically require new or extended ethics approval provided the data was collected ethically and anonymized appropriately, with no new data collection involving human subjects being undertaken.

## Notes

### Competing Interest Statement

The authors have declared no competing interest.

### Funding Statement

The author(s) received no specific funding for this work.

### Author Declarations

This study utilized secondary data collected from the previous STEP project, which was implemented starting in 2018. The initial ethics approval for that project was obtained from the University of South Carolina, covering the period from May 2017 to May 2018, during which data collection took place. The project received approval from different institutions due to the Principal Investigator (PI) shifting institutions during the project. Specifically, the project was reviewed and approved by the institutional review boards of the University of South Carolina (approval period: 5/16/2017 to 5/15/2018), the National Institute of Medical Research of Tanzania (NIMR/HQ/R.8c/Vol.I/1170, approval period: 29th June 2017 to 28th June 2018), and George Washington University in 2021.

